# Sex, age, and hospitalization drive antibody responses in a COVID-19 convalescent plasma donor population

**DOI:** 10.1101/2020.06.26.20139063

**Authors:** Sabra L. Klein, Andrew Pekosz, Han-Sol Park, Rebecca L. Ursin, Janna R. Shapiro, Sarah E. Benner, Kirsten Littlefield, Swetha Kumar, Harnish Mukesh Naik, Michael J. Betenbaugh, Ruchee Shrestha, Annie A. Wu, Robert M. Hughes, Imani Burgess, Patricio Caturegli, Oliver Laeyendecker, Thomas C. Quinn, David Sullivan, Shmuel Shoham, Andrew D. Redd, Evan M. Bloch, Arturo Casadevall, Aaron A.R. Tobian

**Author notes:** corresponding authors: Sabra Klein, Aaron Tobian. co-senior authors (Sabra L. Klein, Evan M. Bloch, Arturo Casadevall, Aaron A.R. Tobian).

## Abstract

Convalescent plasma is currently one of the leading treatments for COVID-19, but there is a paucity of data identifying therapeutic efficacy. A comprehensive analysis of the antibody responses in potential plasma donors and an understanding of the clinical and demographic factors that drive variant antibody responses is needed. Among 126 potential convalescent plasma donors, the humoral immune response was evaluated by a SARS-CoV-2 virus neutralization assay using Vero-E6-TMPRSS2 cells, commercial IgG and IgA ELISA to Spike (S) protein S1 domain (Euroimmun), IgA, IgG and IgM indirect ELISAs to the full-length S or S-receptor binding domain (S-RBD), and an IgG avidity assay. Multiple linear regression and predictive models were utilized to assess the correlations between antibody responses with demographic and clinical characteristics. IgG titers were greater than either IgM or IgA for S1, full length S, and S-RBD in the overall population. Of the 126 plasma samples, 101 (80%) had detectable neutralizing titers. Using neutralization titer as the reference, the sensitivity of the IgG ELISAs ranged between 95-98%, but specificity was only 20-32%. Male sex, older age, and hospitalization with COVID-19 were all consistently associated with increased antibody responses across the serological assays. Neutralizing antibody titers were reduced over time in contrast to overall antibody responses. There was substantial heterogeneity in the antibody response among potential convalescent plasma donors, but sex, age and hospitalization emerged as factors that can be used to identify individuals with a high likelihood of having strong antiviral antibody levels.

**One Sentence Summary:** There is substantial heterogeneity in the antibody response to SARS-CoV-2 infection, with greater antibody responses being associated with male sex, advancing age, and hospitalization with COVID-19.

## Introduction

Severe acute respiratory syndrome coronavirus 2 (SARS-CoV-2), the causative agent of coronavirus disease 2019 (COVID-19), emerged in Wuhan, China in December 2019. Following the rapid, global spread of SARS-CoV-2, in March 2020, COVID-19 was declared a pandemic. To date, over 9 million cases have been confirmed, spanning 188 countries or territories and accounting for over 460,000 deaths (*1*). Preventive and treatment options are limited, of which antibody therapy (i.e. convalescent plasma collected from individuals after recovery from COVID-19) has emerged as a leading treatment for COVID-19 (*2*). Observational findings are encouraging, suggesting improved clinical outcomes in those who are transfused with COVID-19 convalescent plasma (CCP), including radiological resolution, reduction in viral loads, and improved survival (*3-7*). Nonetheless, there is a lack of standardization of units of CCP that are being transfused, in large part due to limited data correlating antibody assays with formal virus neutralization activity.

Antibody responses that target the immunodominant SARS-CoV-2 Spike (S) protein — specifically, those that target the S protein receptor binding domain (S-RBD)— are thought to be highly associated with virus neutralization by blocking the interaction between S-RBD and the virus receptor, angiotensin converting enzyme 2 (AEC2) (*8*). The SARS-CoV-2 S protein is a highly glycosylated, trimeric protein that requires proteolytic processing to become fusogenic and mediate virus-host membrane fusion (*9, 10*). The S-RBD domain is partially masked in the pre-fusion structure of S and must be converted to an “open” conformation for optimal binding of S to ACE2 (*11*). Neutralizing antibodies are of particular interest because they prevent viral infection by blocking cell surface attachment, as well as inhibiting host membrane fusion (*12, 13*). Administration of CCP containing these neutralizing antibodies to individuals with COVID-19 has been shown to result in rapid viral clearance, indicating its functionality as an antiviral agent (*6*). Non-neutralizing antibodies also play a key role in viral clearance as they are needed for antibody dependent cellular cytotoxicity, antibody dependent cell mediated phagocytosis and complement activation (*14*). The contribution of other antibody types such as IgM and IgA to resolution and protection from SARS-CoV-2 infection is not clear. Using plasma samples from 126 recovered COVID-19 patients following mild or moderate disease, we compared commercial ELISAs, two-step Spike protein directed ELISAs, and microneutralization assays, in order to assess how the age and sex of the donor, history of hospitalization for COVID-19, and the time of plasma collection relative to infection could be used to understand the variability of antibody responses to SARS-CoV-2.

## Results

### Immunoglobulin (Ig) isotyping in COVID-19 convalescent plasma

Convalescent plasma was collected from 126 patients with molecular confirmed SARS-CoV-2 infection. The population consisted of more males (56%) than females, with a median age of 42 years (IQR 29-53) (**Table 1**). Most of the patients were classified as having mild to moderate disease, with <10% having been hospitalized with COVID-19. Plasma samples were collected from patients a median of 43 days (IQR 38-48) after an initial PCR+ nasal swab test.

**Table 1.** Demographic data from convalescent plasma donors.

|  | All | Females | Males |
| --- | --- | --- | --- |
| **N (%)** | 126 | 58 (46) | 68 (54) |
| **Age - med (IQR)** | 42 (29 - 53) | 41.5 (28 - 55) | 42 (31.5 - 53) |
| **Age Categories - n (%)** |  |  |  |
| 19-44 | 74 (58.7) | 34 (58.6) | 40 (58.8) |
| 45-64 | 41 (32.5) | 19 (32.8) | 22 (32.4) |
| 65+ | 11 (8.7) | 5 (8.6) | 6 (8.8) |
| **Race/ethnicity - n (%)** |  |  |  |
| White | 94 (74.6) | 42 (72.4) | 52 (76.5) |
| African American | 4 (3.2) | 1 (1.7) | 3 (4.4) |
| Asian | 14 (11.1) | 8 (13.8) | 6 (8.8) |
| Hispanic | 5 (4) | 2 (3.4) | 3 (4.4) |
| Mixed/other/unknown | 9 (7.1) | 5 (8.6) | 4 (5.9) |
| **Hospitalized - n (%)<sup>a</sup>** | 11 (8.9) | 6 (10.7) | 5 (7.4) |
| **No of days - med (IQR)** | 5 (2-6) | 5 (2-5) | 4 (3-6) |
| **Days since swab - med (IQR)** | 43 (38-48) | 44.5 (39-49) | 41 (37-48) |

<sup>a</sup> Hospitalization status missing for 2 donors. Percentages calculated out of total of available data

Plasma samples were used for isotyping antibodies that recognized SARS-COV-2 S antigens. Using the Euroimmun ELISA that recognizes either IgG or IgA against S protein domain S1, we determined that both isotypes were highly detectable in plasma, with arbitrary unit (AU) values of anti-S1 IgG being greater than IgA (*p*<0.05; **Fig. 1A**), but positively associated with each other (r>0.5; **Fig. 1B**). Consistent results were obtained with the indirect ELISAs that recognized either S or S-RBD, in which titers of IgG, as quantified as area under the curve (AUC), were greater than titers of either IgM or IgA (*p*<0.05 in each case; **Fig. 1C** and **1E**). The AUC values for both anti-S and anti-S-RBD IgG were positively associated with the AUC values for anti-S and anti-S-RBD IgM and IgA, respectively (*r*>0.5 in each case; **Fig. 1D** and **1F**). Finally, AUC values for anti-S and anti-S-RBD IgG, IgM, and IgA were correlated with the respective geometric mean titers (**Fig. S1**).

**Fig. 1:**
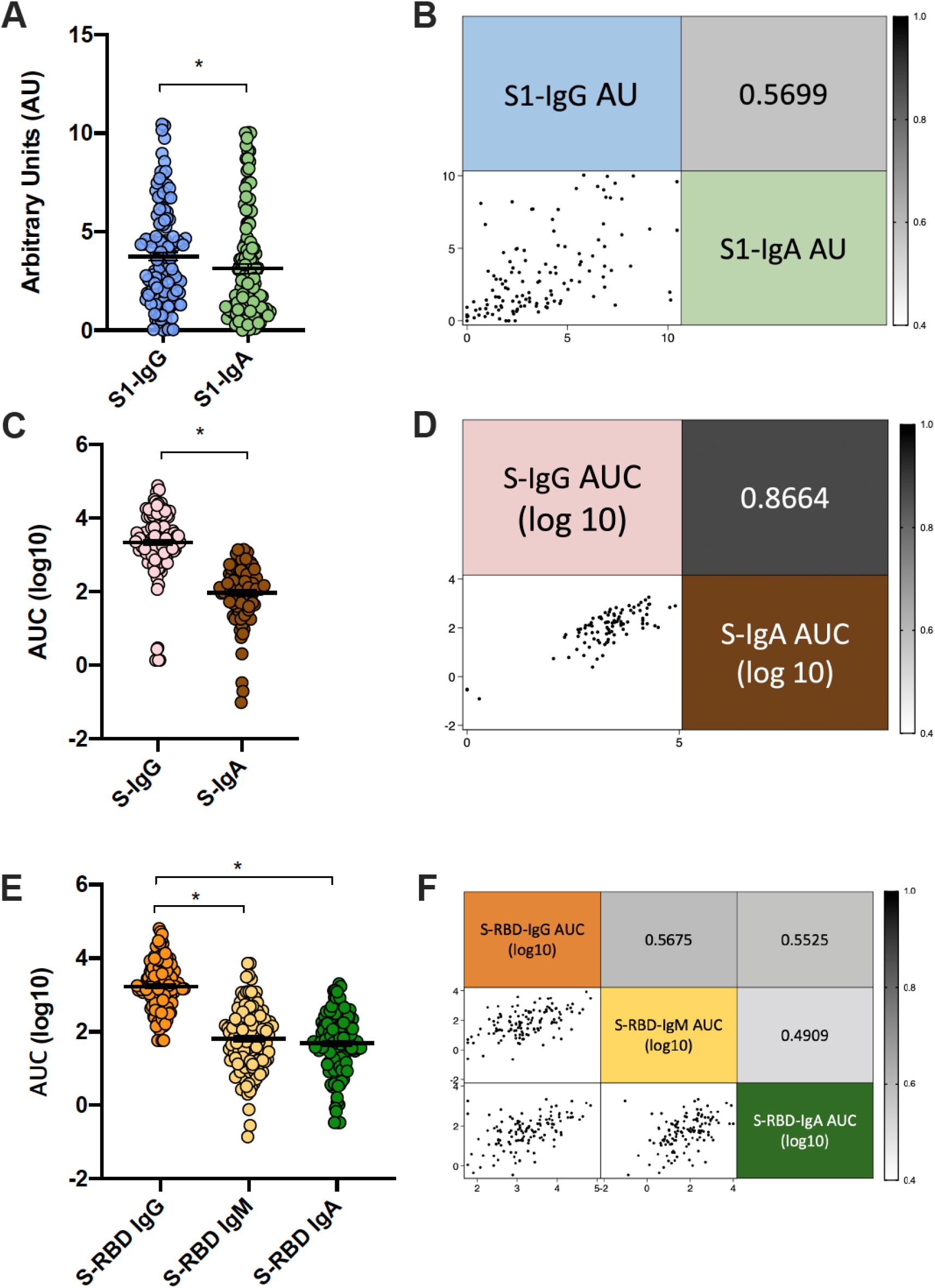
IgG is the primary isotype produced against SARS-CoV-2 spike (S) protein. Convalescent plasma samples from recovered COVID-19 patients were used to assess antibody isotypes that recognize SARS-CoV-2 antigens. Commercial kits from Euroimmun were used to measure total IgG and IgA antibodies against the SARS-CoV-2 Spike (S) protein domain S1 at an optical density of 450nm (OD450) and were compared to a calibrator to yield arbitrary units (AU) (**A**). The correlation between anti-S1 isotypes is graphed, with the r value noted (**B**). Indirect ELISAs were used to measure IgG and IgA antibody levels against S (**C**) and IgG, IgM, and IgA against the S-receptor binding domain (RBD) (**E**) and are graphed as area under the curve (AUC) values. The correlations between IgG and IgA for S (**D**) and IgG, IgM, and IgA for S-RBD (**F**) are included, with r values shown and are shaded darker for higher correlation values or lighter for lower correlation values. Graphs show mean + SEM. (n = 126) \**p* < 0.05 (paired t-test).

### Defining functional antibody in COVID-19 convalescent plasma

To assess the functionality of antibodies that recognize SARS-CoV-2 in convalescent plasma, microneutralization (NT) and IgG avidity assays were performed. The reciprocal plasma dilution providing protection from SARS-CoV-2 was used to calculate the AUC for the NT assay. Of the 126 plasma samples screened, 101 (80%) had detectable NT titers (**Fig. 2A**). The avidity assay defines the binding characteristics of IgG; the OD reading in the presence of various concentrations of urea was used to calculate AU for IgG avidity (**Fig. 2B**). There was a positive correlation of the results from the NT assay with IgG ELISAs for S1, S, and S-RBD and anti-S1 IgG avidity (**Fig. 2C**), with the correlation to S-RBD antibodies being strongest and the anti-S1 IgG avidity being weakest. Because NT titer is currently considered to be the most critical antibody characteristic associated with protection from infection, we assessed the specificity and sensitivity of the three S protein IgG ELISAs against the NT assay avidity (**Fig. 2D**). We designated cutoffs of >20 for NT, < AU 0.8 for S1 IgG, and endpoint titers of <1:320 for S and S-RBD ELISAs. The overall sensitivity of the IgG assays was good, with S1-IgG at 96%, S-IgG at 98%, and S-RBD-IgG at 95%. The specificity of the IgG assays was consistently low, with S1-IgG at 32%, S-IgG at 20%, and S-RBD-IgG at 28%.

**Fig. 2:**
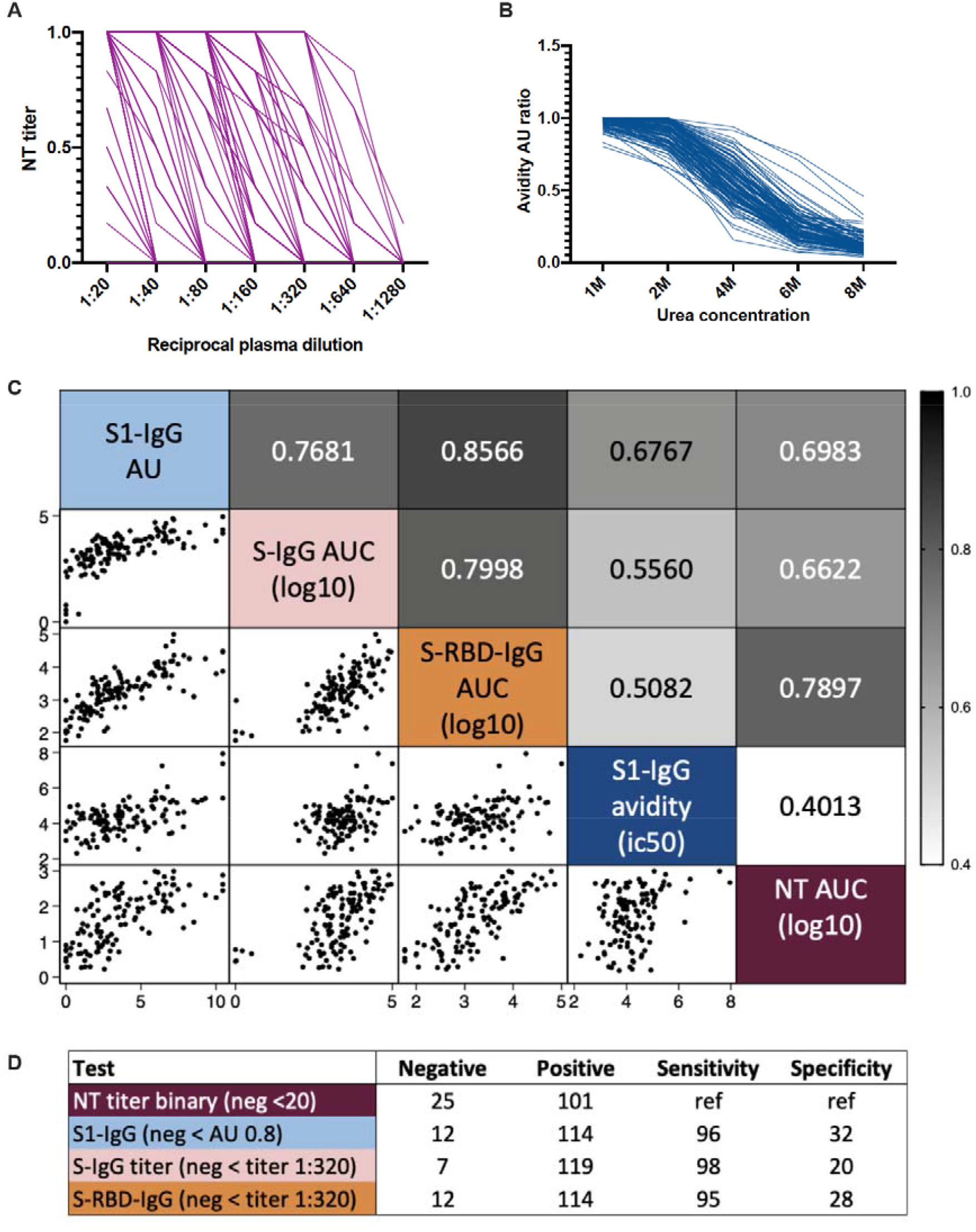
Neutralizing (NT) antibody titers correlate with IgG antibodies that recognize SARS-CoV-2 spike (S) protein. Convalescent plasma samples from recovered COVID-19 patients were used to assess functional antibody levels. Microneutralization assays were performed on each plasma sample in two-fold serial dilutions (**A**). Avidity assays used varying amounts of urea to dissociate the anti-S1 spike protein domain IgG/antigen complex from each plasma sample (represented as arbitrary units, AU) (**B**). The correlation between NT area under the curve (AUC) values, anti-S1 IgG avidity (AU), anti-S1-IgG AU, anti-S-IgG AUC, and anti-S-receptor binding domain (S-RBD)-IgG AUC are shown, with the r values indicated and shaded darker for higher correlation values or lighter for lower correlation values (**C**). For each assay the sensitivity and specificity were defined and compared to the NT AUC, with the negative cutoff value, the number of plasma samples considered positive and negative, as well as how sensitive and specific that assay in reference (ref) to the NT titers (**D**).

### Host factors contributing to improved antibody responses in COVID-19 convalescent plasma

Using the unadjusted NT antibody AUC values, we determined that males consistently had greater neutralizing, anti-S IgG, and anti-RBD IgG than females (*p*<0.05 in each case, **Table S1**). Both NT antibody and anti-RBD IgG titers, in particular, were consistently higher among males than females within diverse age categories and among non-hospitalized patients (**Table S1** and **Fig S2**).

Multiple linear regressions were used to isolate the effects of sex, age, hospitalization, or time since PCR-positive (PCR+) nasal swab on the antibody response to SARS-CoV-2 while adjusting for the other parameters (**Table S2** and **Fig. 3**). As shown in **Fig. 3A-P** and **Table S2**, being male, an older adult, and being hospitalized with COVID-19 were each associated with having greater NT AUC values, anti-S1 IgG AU, anti-S IgG AUC values, and anti-RBD IgG AUC values (*p*<0.05 in each case). In comparing the effect size of each parameter, being hospitalized was associated with the largest increase in antibody response (**Fig. 3Q**). In comparing the four assays, being male, older, and hospitalized had the largest effect on the anti-S1-IgG response. The only antibody measure associated with time (days, scaled by 10) since PCR+ nasal swab test was NT responses, which were reduced as the days since the PCR+ nasal swab test increased (*p*<0.05; **Fig. 3P-Q**).

**Fig. 3:**
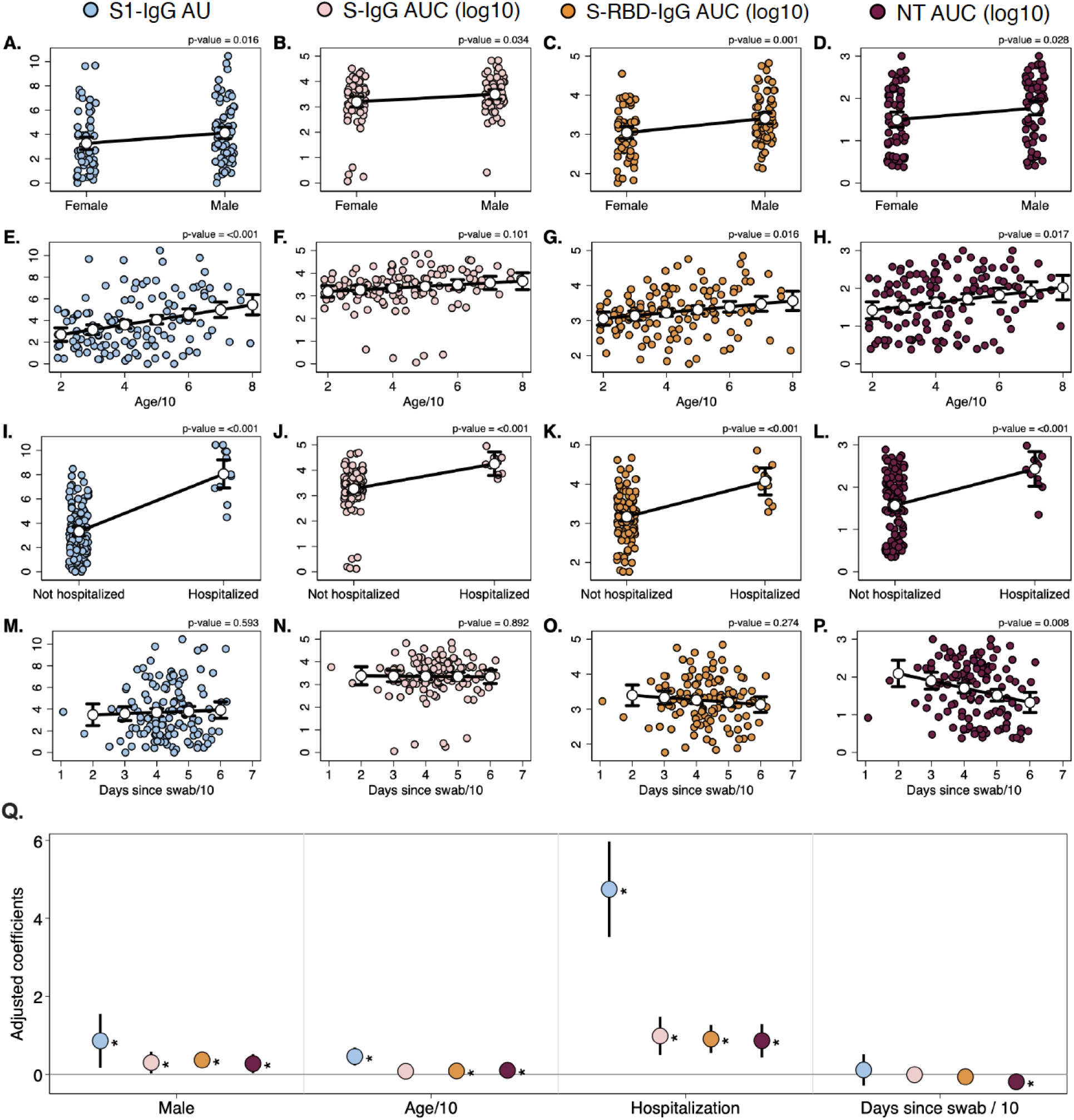
Sex, age, hospitalization, and time since PCR+ nasal swab are associated with antibody responses to SARS-CoV-2. Multiple linear regressions were performed on the continuous outcomes of anti-spike (S) protein domain S1 IgG arbitrary units (AU) (**A, E, I, M**), anti-S-IgG area under the curve (AUC) (**B, F, J, N**), anti-S-RBD AUC (**C, J, K, O**), and neutralizing antibody (NT) AUC (**D, H, L, P**). For each outcome, the model included parameters for the four predictors of interest: sex (**A-D**), age in decades (**E-H**), hospitalization status (**J-L**), and number of days since PCR+ nasal swab (**M-P**). Regression models included the 121 subjects for which complete predictor data was available (date of PCR+ swab was missing for 3 subjects). In each panel, colored circles show the raw data, and white dots show the marginal effect of the given predictor, or the model-predicted outcome (with 95% CI) for the average person for different levels of the given predictor. P-values on top of each panel represent the significance level for the parameter. The four models are summarized in **Q**, where the position of the marker indicates the coefficient value + 95% CI, and stars indicate significance (* = *p*<0.05).

### Predictors of strong antibody responses in donors of COVID-19 convalescent plasma

The convalescent plasma samples were categorized into quartiles based on their neutralizing AUC value, anti-S1 IgG AU, anti-S AUC value, or anti-RBD AUC value resulting in scores ranging from 0 (lowest quartile for each antibody measure) to 12 (highest quartile for each antibody measure) to model the optimal antibody responses in convalescent plasma (**Fig. 4A**). 16/126 (13%) of donors were in the lowest decile in all measured responses.

**Fig. 4:**
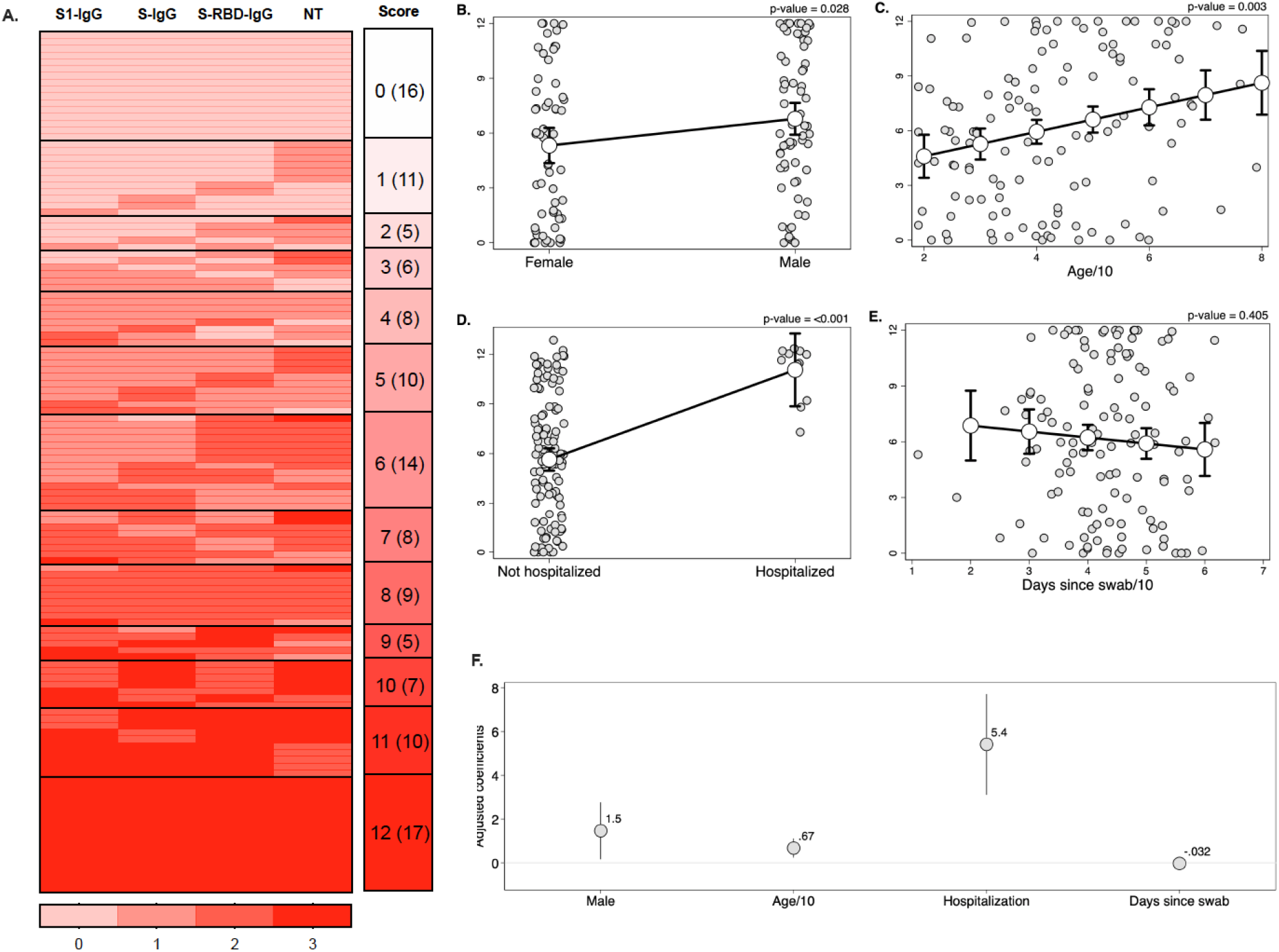
Male sex and hospitalization are predictors of overall greater antibody titers in convalescent plasma. Composite scores were computed for each subject based on the quartile of their response across the anti-spike (S) protein domain S1 IgG, anti-S-IgG, anti-S-receptor binding domain (S-RBD) IgG, and neutralizing antibody (NT) assays (**A**). The distribution of scores among the study population is shown to the right of the heatmap. Multiple linear regression was performed on the continuous outcome of score, including parameters for sex, age in decades, hospitalization status, and number of days since PCR+ nasal swab scaled by ten. For each predictor, the raw data is shown in gray, and the marginal effect + 95% CI of the given predictor for the average individual in the study is shown in white (**B-E**). P-values on top of each panel represent the significance level for the parameter. The model is summarized in **F**, where the position of the marker indicates the coefficient value + 95% CI, or the expected increase in score for a one unit increase in each predictor.

Multiple linear regression on the composite score encompassing the quartiles for each antibody measure revealed that being male, advancing in age, and hospitalized with severe COVID-19 could each predict greater antibody responses against SARS-CoV-2 (**Fig. 4B-D**). In contrast, time elapsed since PCR+ nasal swab was not predictive of greater antibody responses (**Fig. 4E**). In terms of effect size, being male resulted in an average numerical increase in score of 1.5 compared to being female, advancing age by a decade resulted in a <1 numerical increase, and being hospitalized resulted in an average increase of 5 in the quartile score (**Fig. 4F**). Taken together, these data suggest that being hospitalized with severe COVID-19 and male could be used as predictors of greater convalescent plasma antibody responses against SARS-CoV-2.

## Discussion

COVID-19 convalescent plasma has emerged as a leading therapy for hospitalized COVID-19 patients, with thousands of patients treated to date (*5*). There is a compelling argument for why it could be effective either as prophylaxis after exposure, or as treatment for early disease (*15*). Consequently, it is important to measure the antibody response following recovery from infection with SARS-CoV-2 responses to understand characteristics for ideal convalescent plasma donation. These data suggest that diverse isotypes of antibody are detectable in plasma approximately 40 days following a positive PCR+ test for SARS-CoV-2, and IgG is the prominent isotype across diverse assays and analyses. Although the commercial ELISA to S1 protein and ELISAs to S and S-RBD correlate with the neutralizing titers, they have poor specificity. The poor specificity of the IgG ELISAs with NT titers suggest that overall antibody levels may not accurately reflect NT activity in plasma. In addition, while overall antibody levels seemed constant, there was a significant decrease in NT antibody titers over time. Overall, greater NT and IgG titers were associated with male sex, older age, and a history of hospitalization, but further investigation is needed to determine if common or divergent factors are driving these associations.

The heterogeneity in the antibody response demonstrated in this study is consistent with previously published data. While reports from China suggest that the majority of individuals generate greater titers of antibodies ≥ 14 days after resolution of symptoms (*16*), 30% of patients do not appear to develop sufficient neutralizing antibody titers following infection (*17*). In the present study, 20% of individuals had no detectable neutralizing antibody. Male sex, advancing age, and hospitalization with severe COVID-19 were associated with greater NT and IgG responses to SARS-CoV-2. Greater IgG titers were correlated with worse COVID-19 outcomes, which is also reflected in the link between greater titers and increased age (*18*).

Male sex also is associated with greater risk of more severe COVID-19 outcomes (*19*). The greater antibody responses in convalescent plasma from males as compared with females is striking given that females usually mount stronger immune responses than males (*20*). One possible explanation for this apparent reversal in sex-related differences in antibody responses to SARS-CoV-2 is that males with COVID-19 tend to have more severe disease than females, and enhanced inflammatory responses associated with increased disease severity could drive higher B cell recruitment and consequently, more antibody production. In this regard, the magnitude of antibody responses also correlates with disease severity in other infectious diseases, such as active tuberculosis (*21*).

There are limitations associated with this study. The samples were cross-sectional with a relatively tight window of collection. Therefore, the kinetics of the complete antibody response over time could not be determined, and it was difficult to assess how the time relative to the initial diagnosis correlates with the overall titer. The sampled population, however, represented a clinically diverse population with a wide age range that is representative of the blood donor population. The study was also limited by the lack of measurement of non-direct measures of antibody function (e.g., phagocytosis, antibody-dependent cellular cytotoxicity), but the importance of these mechanisms is not known.

Initially, the FDA recommended that convalescent plasma donors would optimally have ELISA titers exceeding 1:320; this was subsequently lowered given concerns that insufficient donors would attain this threshold (*15*). Currently, the FDA recommends a NT concentration of >160, yet allow for a lower titer (1:80) if an alternative is unavailable (*22*). The FDA, however, has not been prescriptive about the assays used to derive these titer levels despite the potential variability by assay. Data from the Expanded Access Program and clinical trials are urgently needed to interpret the titers with respect to that clinical outcomes and prevention. These results provide a roadmap to select individuals who are likely to have high levels of neutralizing and anti-SARS-CoV-2 IgG antibodies to be preferred convalescent plasma donors.

## Materials and Methods

### Study participants, blood sample processing, and storage

Individuals with a history of COVID-19 who were interested in donating convalescent plasma were contacted by study personnel. All subjects had to be at least 18 years old and have had a confirmed diagnosis of SARS-CoV-2 by detectable RNA on a nasopharyngeal swab. Donors were informed that they needed to satisfy standard eligibility criteria for blood donation (e.g., not pregnant within the last six weeks, never been diagnosed or have risk factors for transfusion-transmitted infections such as HIV, hepatitis B virus or hepatitis C virus). These individuals were then invited to participate in the study. Basic demographic information (age, sex, and hospitalization with COVID-19) was obtained from the subject (i.e. potential donor); confirmation of the original diagnosis of SARS-CoV-2 was required either by medical chart review or sharing of source documentation, including the date the diagnosis was ascertained. Eligible subjects were enrolled in the study under full informed consent; following consent, ∼25 mL of whole blood was collected in ACD tubes. The samples were separated into plasma and peripheral blood mononuclear cells within 12 hours of collection. The plasma samples were immediately frozen at −80°C. The Johns Hopkins University School of Medicine Institutional Review Board reviewed and approved the sample collection and overall study.

### Plasmid preparation

Recombinant plasmid constructs containing modified Spike (S) protein or S protein Receptor Binding Domain (RBD) and a beta-lactamase (amp) gene were obtained (*23*) and amplified in E.coli after transformation and growth on LB agar plates coated with Ampicillin. The plasmids were extracted using GigaPrep kits (Thermo Fisher Scientific) and eluted in molecular biology grade water.

### Recombinant protein expression

HEK293.2sus cells (ATCC) were obtained and adapted to Freestyle™ F-17 medium (Thermo Fisher Scientific) and BalanCD® (Irvine Scientific) using polycarbonate shake flasks (Fisherbrand) with 4mM GlutaMAX supplementation (Thermo Fisher Scientific). The cells were routinely maintained every 4 days at a seeding density of 0.5 million cells/mL. They were cultured at 37°C, 90% humidity with 5% CO2 for cells in BalanCD® while those in F-17 were maintained at 8% CO2. Cells were counted using trypan blue dye (Gibco) exclusion method and a haemocytometer. Cell viability was always maintained above 90%. Twenty-four hours prior to transfection (Day −1), the cells were seeded at a density of 1 million cells/mL, ensuring that the cell viability was above 90%. Polyethylenimine (PEI) stocks, with 25 kDa molecular weight (Polysciences), were prepared in MilliQ water at a concentration of 1 mg/mL. This was sterile filtered through a 0.22 μm syringe filter (Corning), aliquoted and stored at −20°C.

On the day of transfection (Day 0), the cells were counted to ensure sufficient growth and viability. OptiPRO™ SFM (Gibco) was used as the medium for transfection mixture. For 100 mL of cell culture, 2 tubes were aliquoted with 6.7 mL each of OptiPRO™, one for PEI and the other for rDNA. DNA:PEI ratio of 1:3.5 was used for transfection. A volume of 350 μl of prepared PEI stock solution was added to tube 1 while 100 μg of rDNA was added to tube 2 and incubated for 5 minutes. Post incubation, these were mixed together, incubated for 10 minutes at RT and then added to the culture through gravity addition. The cells were returned back to the 37°C incubator. A day after transfection (Day 1), the cells were spun down at 1,000 rpm for 7 minutes at RT and resuspended in fresh media with GlutaMAX™ supplementation. 3-5 hours after resuspension, 0.22 μm sterile filtered Sodium butyrate (EMD Millipore) was added to the flask at a final concentration of 5 mM (Grünberg et al.). The cells were allowed to grow for a period of 4-5 days. Cell counts, viability, glucose and lactate values were measured every day. Cells were harvested when either the viability fell below 60% or when the glucose was depleted, by centrifugation at 5000 rpm for 10 minutes at RT. Cell culture supernatants containing either recombinant RBD or S protein were filtered through 0.22 μm PES membrane stericup filters (Millipore Sigma) to remove cell debris and stored at −20°C until purification.

### Protein purification

Protein purification by immobilized metal affinity chromatography (IMAC) and gravity flow was adapted from previous methods (*23*). After washing with Phosphate-Buffered Saline (PBS; Thermo Fisher Scientific), Nickel-Nitrilotriacetic acid (Ni-NTA) agarose (Qiagen) was added to culture supernatant followed by overnight incubation (12-16 hours) at 4 °C on a rotator. For every 150 mL of culture supernatant, 2.5 mL of Ni-NTA agarose was added. 5mL gravity flow polypropylene columns (Qiagen) were equilibrated with PBS. One polypropylene column was used for every 150 mL of culture supernatant. The supernatant-agarose mixture was then loaded onto the column to retain the agarose beads with recombinant proteins bound to the beads. Each column was then washed, first with 1X culture supernatant volume of PBS and then with 25 mL of 20 mM imidazole (Millipore Sigma) in PBS wash buffer to remove host cell proteins. Recombinant proteins were then eluted from each column in three fractions with 5 mL of 250 mM imidazole in PBS elution buffer per fraction giving a total of 15 mL eluate per column. The eluate was subsequently dialyzed several times against PBS using Amicon Ultra Centrifugal Filters (Millipore Sigma) at 7000 rpm for 20 minutes at 10 °C to remove the imidazole and concentrate the eluate. Filters with a 10 kDa molecular weight cut-off were used for RBD eluate whereas filters with a 50 kDa molecular weight cut-off were used for full length S eluate. The final concentration of the recombinant RBD and S proteins was measured by bicinchoninic acid (BCA) assay (Thermo Fisher Scientific), and purity was assessed on 10% SDS-PAGE (Bio-Rad) followed by Coomassie blue staining. After sufficient destaining in water overnight, clear single bands were visible for RBD and S proteins at their respective molecular sizes.

### Viruses and cells

Vero-E6 cells (ATCC CRL-1586) and Vero-E6-TMPRSS2 cells (*24*) were cultured in Dulbecco’s modified Eagle medium (DMEMD) containing 10% fetal bovine serum (Gibco), 1 mM glutamine (Invitrogen), 1 mM sodium pyruvate (Invitrogen), 100 U/ml of penicillin (Invitrogen), and 100 μg/ml of streptomycin (Invitrogen) (complete media or CM). Cells were incubated in a 5% CO2 humidified incubator at 37°C.

The SARS-CoV-2/USA-WA1/2020 virus was obtained from BEI Resources. The infectious virus titer was determined on Vero cells using a 50% tissue culture infectious dose (TCID50) assay as previously described for SARS-CoV (*25, 26*). Serial 10-fold dilutions of the virus stock were made in infection media (IM, which is identical to CM except the FBS is reduced to 2.5%), then then 100 μl of each dilution was added to Vero cells in a 96-well plate in sextuplicate. The cells were incubated at 37°C for 4 days, visualized by staining with naphthol blue-black, and scored visually for cytopathic effect. A Reed and Muench calculation was used to determine TCID50 per ml (*27*).

### Enzyme-linked Immunosorbent Assays (ELISAs)

#### Commercial ELISAs and Avidity

The Euroimmun Anti-SARS-CoV-2 ELISA (Mountain Lakes, NJ) for both IgA and IgG was validated in a Clinical Laboratory Improvement Amendments (CLIA)-certified laboratory. The assay was performed per the manufacturer’s specification. The optical density (OD) of the sample divided by the OD of the calibrator from that run, and the ratio is the arbitrary unit (AU). Per the manufacturer, an AU 0-0.79 was considered negative, 0.80-0.99 was borderline and ≥ 1.0 was positive.

To measure anti-SARS-CoV-2 IgG avidity, each reaction utilized the following components: 100 μl of plasma (1:101 dilution per manufactures protocol), 100 μl of undiluted positive, negative and calibrator controls. Plates containing reaction components were incubated for 1 hour at 37°C followed by 3 washes. A 300 μl volume of wash buffer containing urea at varying concentrations (0M, 1M, 2M, 4M, 6M or 8M) were added to the plates and incubated at 37°C for 10 minutes (*28*). Plates were washed 3 times, followed by the manufacturer’s protocol for addition of conjugate and substrate. Ratios of ≥ 0.8 were considered positive. DC50 (Dissociation Constant 50) calculations were performed using AAT Bioquest IC50 calculator using four parameter logistic regression model (AAT Bioquest, Inc. (2020, June 09). *Quest Graph™ IC50 Calculator was* retrieved from https://www.aatbio.com/tools/ic50-calculator.

#### Indirect ELISAs

The protocol was adapted from a published protocol from Dr. Florian Krammer’s laboratory (*23*). Ninety-six well plates (Immulon 4HBX, Thermo Fisher) were coated with either full length S protein or S-RBD) at a volume of 50 μl of 2 μg/ml of diluted antigen in filtered, sterile 1xPBS (Thermo Fisher) at 4°C overnight. Coating buffer was removed, plates were washed three times with 300 μl of PBS-T wash buffer (1xPBS plus 0.1% Tween 20, Fisher Scientific), and blocked with 200 μl of PBS-T with 3% non-fat milk (milk powder, American Bio) by volume for one hour at room temperature. All plasma samples were heat inactivated at 56°Con a heating block for one hour prior to use. Negative control samples were prepared at 1:10 dilutions in PBS-T in 1% non-fat milk and plated at a final concentration of 1:100. A monoclonal antibody (mAb) towards the SARS-CoV-2 spike protein was used as a positive control (1:5,000, Sino Biological). For serial dilutions of plasma on either S or S-RBD-coated plates, plasma samples were prepared in three-fold serial dilutions starting at 1:20 in PBS-T in 1% non-fat-milk. Blocking solution was removed and 10 μl of diluted plasma was added in duplicates to plates and incubated at room temperature for two hours. Plates were washed three times with PBS-T wash buffer and 50 μl secondary antibody was added to plates and incubated at room temperature for one hour. Anti-human secondary antibodies used included Fc-specific total IgG HRP (1:5,000 dilution, Invitrogen), IgM heavy chain HRP (1:5,000, Invitrogen), and IgA cross-adsorbed HRP (1:5,000, Invitrogen); all were prepared in PBS-T plus 1% non-fat milk. Plates were washed and all residual liquid removed before adding 100 μl of SIGMAFAST OPD (o-phenylenediamine dihydrochloride) solution (Sigma Aldrich) to each well, followed by incubation in darkness at room temperature for ten minutes. To stop the reaction, 50 μl of 3M hydrochloric acid (HCl, Fisher Scientific) was added to each well. The OD of each plate was read at 490nm (OD_490_) on a SpectraMax i3 ELISA plate reader (BioTek). The positive cutoff value for each plate was calculated by summing the average of the negative values and three times the standard deviation of the negatives. All values at or above the cutoff value were considered positive.

### Microneutralization assay

Plasma neutralization titers were determined as described for SARS-CoV (*29*). Two-fold dilutions of plasma (starting at a 1:20 dilution) were made in IM. Infectious virus was added to the plasma dilutions at a final concentration of 1×10^4^ TCID50/ml (100 TCID50 per 100ul). The samples were incubated for one hour at room temperature, then 100 uL of each dilution was added to one well of a 96 well plate of VeroE6-TMPRSS2 cells in sextuplet for 6 hours at 37°C. The inoculums were removed, fresh IM was added, and the plates were incubated at 37°C for 2 days. The cells were fixed by the addition of 150 uL of 4% formaldehyde per well, incubated for at least 4 hours at room temperature, then stained with Napthol blue black. The nAb titer was calculated as the highest serum dilution that eliminated cytopathic effect (CPE) in 50% of the wells.

### Statistical analyses

#### Descriptive analyses

Area under the curve (AUC) values were computed by plotting normalized OD values against sample dilution for ELISAs. AUC for NT assays utilized the exact number of wells protected from infection at each plasma dilution. For each assay, samples with titers below the limit of detection were assigned an arbitrary AUC value of half of the lowest measured AUC value. The data were then log-transformed to achieve a normal distribution. Descriptive statistics stratified by sex were presented as medians and interquartile ranges, and male-female comparisons overall and in each age category were done using simple T-tests. AUC values for IgG, IgA and IgM were compared using a one-way ANOVA. Correlations between antibody isotypes and assays were assessed using Pearson’s correlation coefficient. Where binary cut-offs were available, IgG data were dichotomized using the 1:320 cut-off originally recommended by the FDA (*15*) or the cut-off of AU > 0.8 suggested by the manufacturer. Sensitivity and specificity were then calculated using neutralizing antibody (i.e. titer > 1:20) as the reference assay.

#### Predictors of assay-specific responses

Multiple linear regressions were performed to assess the impact of the demographic (age in decades and sex) and clinical factors (hospitalization status and days since PCR+ swab scaled by 10) on S1-IgG OD values, log AUC values for anti-RBD and anti-spike IgG, as well as neutralizing antibody. All predictor estimates were adjusted for the three other parameters in the model. Various additional parameters were tested, including and interaction term between age and sex and linear splines at different ages, but decreased the overall fit of the model and were therefore not included in further analysis. Data are presented as the marginal effect of each predictor for the average person in the study population (*30*) along with coefficients and 95% confidence intervals of each estimate.

#### Composite score representing overall quality of antibody response

Composite scores were computed to provide a single metric as a proxy for the overall quality of the antibody response. The responses for S1-IgG, S-IgG, S-RBD and neutralizing assays were divided into quartiles, and subjects were assigned a score of 0 (lowest quartile) to 3 (highest) quartile for each assay. The assay-specific scores were summed to create the composite score, ranging from 0 (lowest quartile for each assay) to 12 (highest quartile for each assay). A multiple linear regression model was then performed on the composite score, including parameters for sex, age in decades, hospitalization status and number of days since PCR+ swab (scaled by ten). As above, data are presented as the marginal effect of each predictor for the average person in the study population (*30*) along with coefficients and 95% confidence intervals of each estimate. All analyses were performed using GraphPad Prism 8 and Stata 15.

## Data Availability

All data are contained in the manuscript.

## Acknowledgments

We are grateful for all of the study participants who donated plasma, the clinical staff, including Sonali Thapa and Liz Martinez, Mary De’Jarnette, Carlos Aguado, Peggy Iraola, Jackie Lobien who collected samples, and the technical staff, including Yolanda Eby, Rey Fernandez, Haley Schmidt, Charles Kirby, Ethan Klock, Owen Baker, Jernelle Miller, and Morgan Keruly who aliquoted and stored samples for this study. We thank Florian Krammer of the Icahn School of Medicine at Mount Sinai for providing protocols, plasmids, and initial stocks of ELISA antigens.

## Funding

This work was supported in part by NIH Specialized Center of Research Excellence U54AG062333 (S.L.K, A.P., H-S.P, J.S.); NIH Center of Excellence in Influenza Research and Surveillance HHSN272201400007C (A.P., K.L., S.L.K., R.L.U.); T32A1007417 Molecular and Cellular Basis of Infectious Diseases (R.L.U.); National Institute of Allergy and Infectious Diseases (NIAID) AI052733 and AI15207 (A.C.); NIAID R01AI120938 and R01AI128779 (A.A.R.T); the Division of Intramural Research, NIAID (O.L., T.Q.); National Heart Lung and Blood Institute 1K23HL151826-01 (E.B.M) and HL059842 (A.C.). Bloomberg Philanthropies (A.C.); Department of Defense W911QY2090012 (A.C. and D.S.).

## Author contributions

E.M.B., A.C., and A.T. conceived and designed the study; E.M.B. and A.T. wrote the IRB protocol; R.S., A.W, R.M.H., I.B., E.M.B. and A.T recruited participants; S.L.K., A.P., H-S.P., R.L.U., K.L., O.L., T.Q. S.E.B., A.R., E.M.B, and A.T. carried out all experiments; H.M.N., S.K. and M.J.B. produced recombinant SARS-CoV-2 proteins; J.R.S. performed statistical analyses; S.L.K., A.P., H-S.P., R.L.U., J.S., E.M.B., A.C., and A.T. wrote the manuscript with substantial input from all co-authors.

## Competing interests

EMB reports personal fees and non-financial support from Terumo BCT, personal fees and non-financial support from Grifols Diagnostics Solutions, outside of the submitted work; EMB is a member of the United States Food and Drug Administration (FDA) Blood Products Advisory Committee. Any views or opinions that are expressed in this manuscript are that of the author’s, based on his own scientific expertise and professional judgment; they do not necessarily represent the views of either the Blood Products Advisory Committee or the formal position of FDA, and also do not bind or otherwise obligate or commit either Advisory Committee or the Agency to the views expressed.

## Data and materials availability

All data are contained in the manuscript.

## Supplementary Materials

**Fig. S1:**
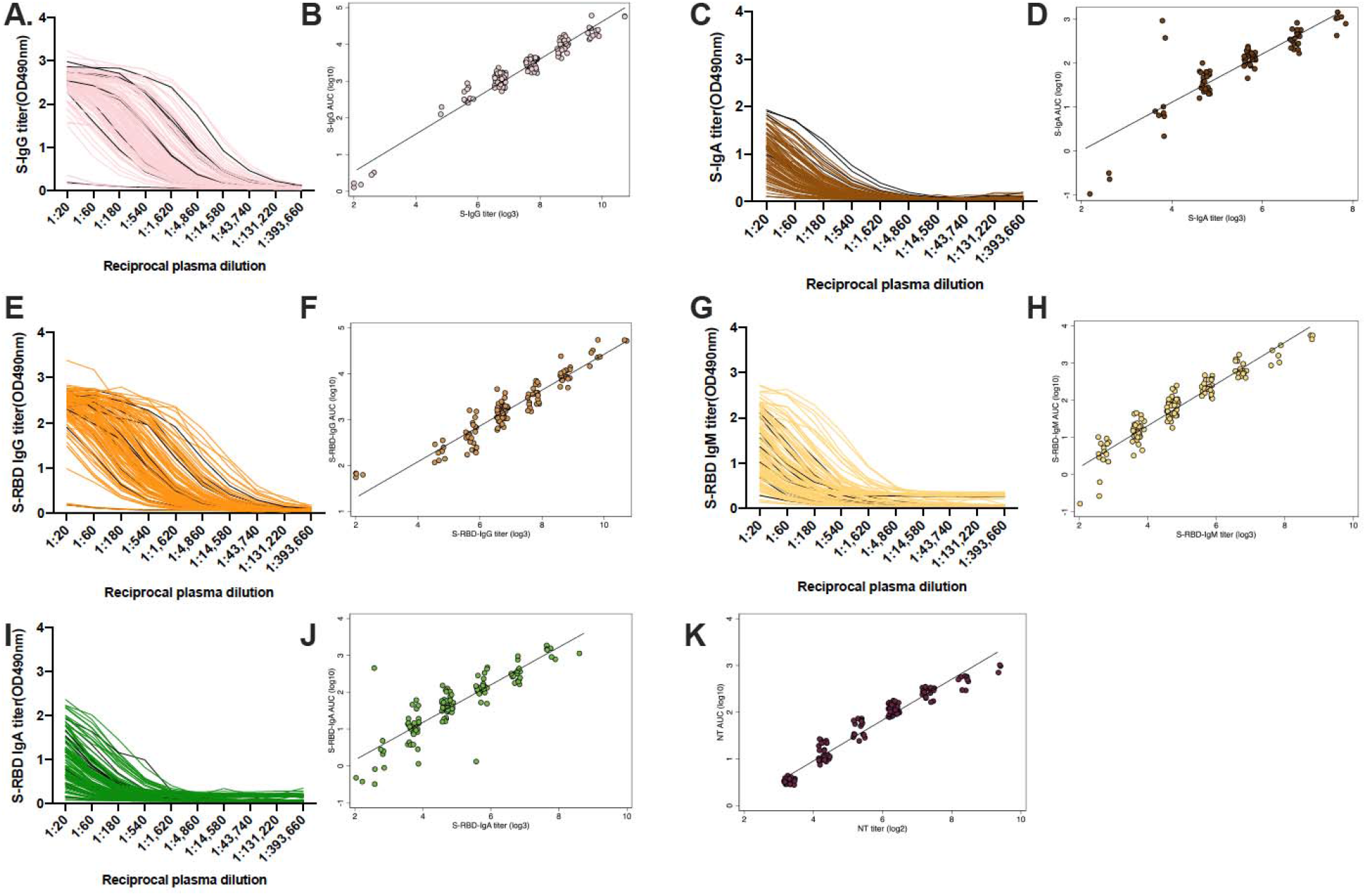
Reciprocal antibody titers and area under the curve comparisons for quantification of anti-spike (S) and anti-S-receptor binding domain (S-RBD) IgG, IgM, and IgA. The optical density at 490nm for each three-fold, serially diluted plasma sample was measured in the indirect ELISAs. ELISAs measuring IgG and IgA in serial dilution against the spike (S) protein (**A, C**) and IgG, IgM, and IgA against the S-receptor binding domain (S-RBD) are shown, with hospitalized patients represented by black lines (**E, G, I**). The correlation between the antibody titer and the area under the curve (AUC) are shown for IgG and IgA against S (**B, D**) and for IgG, IgM, and IgA against S-RBD (**F, H, J**). The correlation between the neutralizing antibody (NT) titer and the calculated AUC is shown (**K**).

**Fig. S2.**
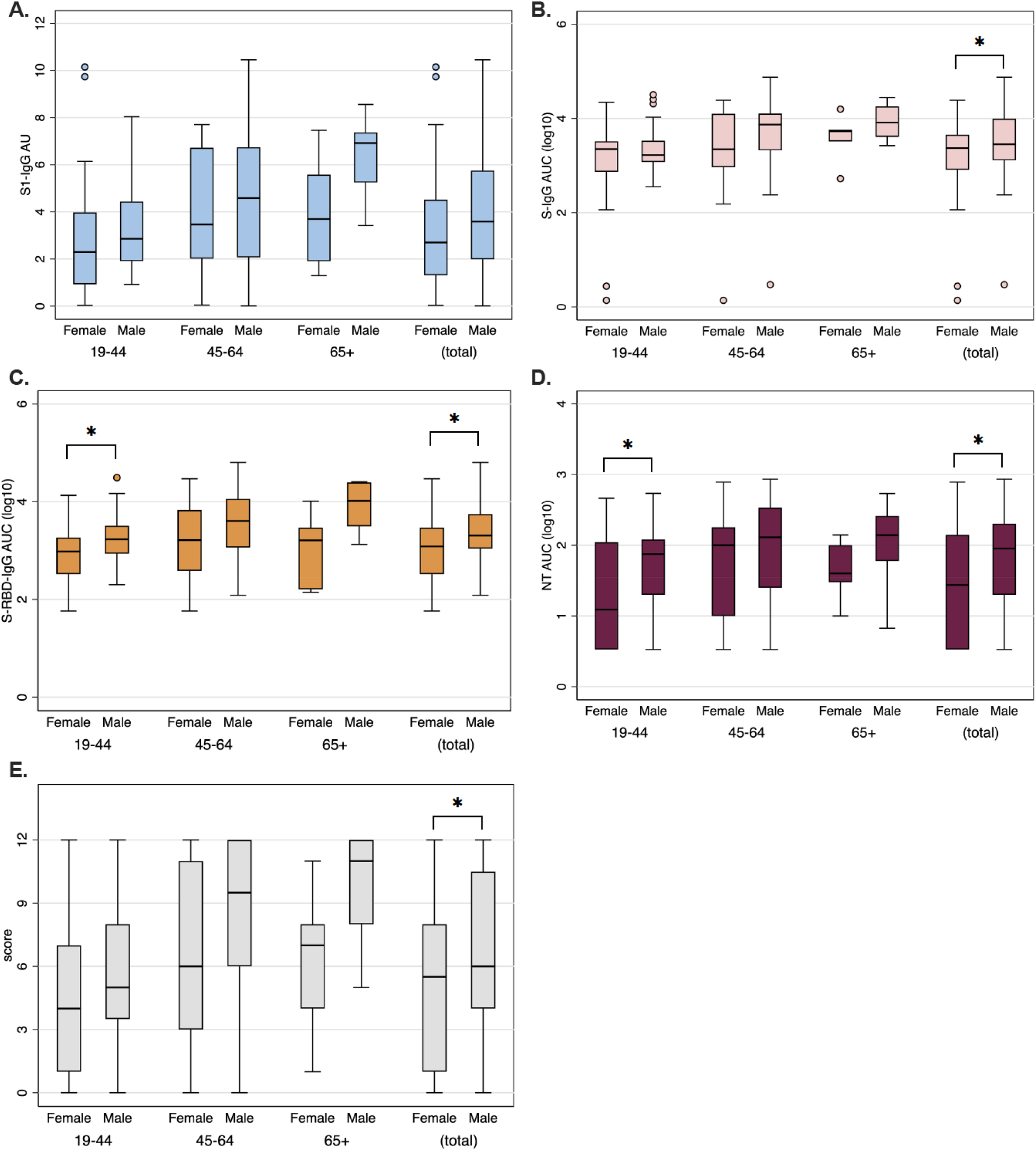
Unadjusted boxplots showing age-associated antibody responses for male and female convalescent plasma donors. Box plots by sex and age categories are shown for anti-spike (S) protein domain S1 IgG arbitrary units (AU) (**A**), anti-S-IgG area under the curve (AUC) (**B**), anti-S-receptor binding domain (S-RBD) AUC (**C**), and neutralizing antibody (NT) AUC (**D**). Sex differences in each age category were analyzed using simple t-tests, and significance is indicated on top of each comparison where appropriate (* = *p*-value <0.05).

**Table S1.** Unadjusted neutralizing (NT) area under the curve (AUC) values, anti-spike (S) protein domain S1 IgG arbitrary units (AU), anti-S IgG AUC values, and anti-S-receptor binding domain (S-RBD) IgG AUC values for all 126 patients and broken down by sex.

| Med (IQR) | All (N = 126) | Females (N = 58) | Males (N = 68) | Sex p-value |
| --- | --- | --- | --- | --- |
| **NT AUC (log10)** |  |  |  |  |
| **Sex** | 1.78 (1-1.78) | 1.44 (.53-2.14) | 1.95 (1.3-2.3) | **0.023** |
| **Age Categories** |  |  |  |  |
| 19-44 | 1.57 (1-2.08) | 1.09 (.53-2.04) | 1.87 (1.3-2.08) | **0.043** |
| 45-64 | 2.04 (1.3-2.48) | 2 (1-2.25) | 2.11 (1.4-2.53) | 0.383 |
| 65+ | 2 (1.48-2.2) | 1.6 (1.48-2) | 2.14 (1.78-2.41) | 0.330 |
| **Hospitalized** |  |  |  |  |
| No | 1.74 (1-2.14) | 1.33 (.53-2.08) | 1.95 (1.3-2.2) | **0.012** |
| Yes | 2.48 (2-2.73) | 2.39 (2-2.5) | 2.73 (2.08-2.73) | 0.790 |
| **S1-IgG AU** |  |  |  |  |
| **Sex** | 3.15 (1.79-3.15) | 2.7 (1.31-4.52) | 3.59 (1.99-5.75) | 0.067 |
| **Age Categories** |  |  |  |  |
| 19-44 | 2.75 (1.6-4.33) | 2.29 (.93-3.97) | 2.86 (1.91-4.44) | 0.321 |
| 45-64 | 4.36 (2.04-6.72) | 3.47 (2.02-6.72) | 4.58 (2.07-6.75) | 0.283 |
| 65+ | 5.58 (3.42-7.37) | 3.7 (1.91-5.58) | 6.92 (5.25-7.37) | 0.100 |
| **Hospitalized** |  |  |  |  |
| No | 2.90 (1.79-4.54) | 2.5 (1.31-3.97) | 3.19 (1.95-4.66) | **0.026** |
| Yes | 8.04 (6.97-10.15) | 7.45 (6.97-9.74) | 8.97 (8.04-10.38) | 0.429 |
| **S-IgG AUC (log10)** |  |  |  |  |
| **Sex** | 3.41 (3.05-3.41) | 3.37 (2.91-3.65) | 3.45 (3.11-3.99) | **0.034** |
| **Age Categories** |  |  |  |  |
| 19-44 | 3.27 (3.03-3.51) | 3.35 (2.87-3.51) | 3.22 (3.07-3.53) | 0.180 |
| 45-64 | 3.53 (3.16-4.1) | 3.34 (2.97-4.1) | 3.87 (3.32-4.1) | 0.156 |
| 65+ | 3.76 (3.51-4.2) | 3.73 (3.51-3.76) | 3.91 (3.61-4.25) | 0.257 |
| **Hospitalized** |  |  |  |  |
| No | 3.35 (3.05-3.61) | 3.34 (2.91-3.51) | 3.36 (3.09-3.84) | **0.044** |
| Yes | 4.24 (3.9-4.5) | 4.21 (3.78-4.34) | 4.5 (4.18-4.71) | 0.143 |
| **S-RBD-IgG AUC (log10)** |  |  |  |  |
| **Sex** | 3.21 (2.79-3.21) | 3.09 (2.52-3.47) | 3.31 (3.04-3.75) | **0.001** |
| **Age Categories** |  |  |  |  |
| 19-44 | 3.15 (2.75-3.46) | 2.98 (2.52-3.27) | 3.23 (2.94-3.51) | **0.020** |
| 45-64 | 3.32 (3.05-3.93) | 3.21 (2.59-3.83) | 3.61 (3.06-4.06) | 0.097 |
| 65+ | 3.5 (3.13-4.04) | 3.21 (2.21-3.47) | 4.02 (3.5-4.4) | 0.051 |
| **Hospitalized** |  |  |  |  |
| No | 3.16 (2.77-3.58) | 3.04 (2.52-3.37) | 3.27 (2.99-3.69) | **0.001** |
| Yes | 4 (3.59-4.47) | 3.92 (3.38-4.13) | 4.32 (4-4.49) | 0.189 |
| **Composite score** |  |  |  |  |
| **Sex** | 6 (2-6) | 5.5 (1-8) | 6 (4-10.5) | **0.032** |
| **Age Categories** |  |  |  |  |
| 19-44 | 5 (2-8) | 4 (1-7) | 5 (3.5-8) | 0.143 |
| 45-64 | 8 (3-11) | 6 (3-11) | 9.5 (6-12) | 0.281 |
| 65+ | 8 (5-12) | 7 (4-8) | 11 (8-12) | 0.105 |
| **Hospitalized** |  |  |  |  |
| No | 6 (2-9) | 4.5 (1-8) | 6 (4-10) | **0.021** |
| Yes | 12 (9-12) | 12 (9-12) | 12 (11-12) | 0.642 |

**Table S2.** Linear regression model coefficients (95% CI) for neutralizing (NT) area under the curve (AUC) values, anti-spike (S) protein domain S1 IgG arbitrary units (AU), anti-S IgG AUC values, and anti-S-receptor binding domain (S-RBD) IgG AUC values for all 126 patients and broken down by sex, age, hospitalization, and days since PCR+ nasal swab.

|  | Coefficient | 95% CI | p-value |
| --- | --- | --- | --- |
| **NT AUC (log10)** |  |  |  |
| Male | 0.273 | (.03, .516) | **0.028** |
| Age/10 | 0.1 | (.018, .181) | **0.017** |
| Hospitalization | 0.862 | (.433, 1.291) | **<0.001** |
| Days since swab / 10 | -0.192 | (-.334, -.05) | **0.008** |
| **S1-IgG AU** |  |  |  |
| Male | 0.857 | (.163, 1.55) | **0.016** |
| Age/10 | 0.457 | (.225, .689) | **<0.001** |
| Hospitalization | 4.746 | (3.522, 5.971) | **<0.001** |
| Days since swab / 10 | 0.109 | (-.295, .514) | 0.593 |
| **S-IgG AUC (log10)** |  |  |  |
| Male | 0.299 | (.023, .575) | **0.034** |
| Age/10 | 0.077 | (-.015, .169) | 0.101 |
| Hospitalization | 0.982 | (.495, 1.469) | **<0.001** |
| Days since swab / 10 | -0.011 | (-.172, .15) | 0.892 |
| **S-RBD-IgG AUC (log10)** |  |  |  |
| Male | 0.365 | (.16, .57) | **0.001** |
| Age/10 | 0.084 | (.016, .153) | **0.016** |
| Hospitalization | 0.905 | (.543, 1.267) | **<0.001** |
| Days since swab / 10 | -0.066 | (-.186, .053) | 0.274 |
| **Composite score** |  |  |  |
| Male | 1.462 | (.157, 2.766) | **0.028** |
| Age/10 | 0.672 | (.236, 1.108) | **0.003** |
| Hospitalization | 5.411 | (3.108, 7.714) | **<0.001** |
| Days since swab / 10 | -0.321 | (-1.082, .439) | 0.405 |

Estimates adjusted for all other predictors in table.
Coefficients represent increase in AUC, AU or score for a one unit increase in predictor.

## Notes

### Author Declarations

The Johns Hopkins University School of Medicine Institutional Review Board reviewed and approved the sample collection and overall study.

## References

1. WHO, WHO, Ed. (2020), vol. 2020.

2. Casadevall, L. A. Pirofski, The convalescent sera option for containing COVID-19. J Clin Invest, (2020).

3. C. Shen, Z. Wang, F. Zhao, Y. Yang, J. Li, J. Yuan, F. Wang, D. Li, M. Yang, L. Xing, J. Wei, H. Xiao, Y. Yang, J. Qu, L. Qing, L. Chen, Z. Xu, L. Peng, Y. Li, H. Zheng, F. Chen, K. Huang, Y. Jiang, D. Liu, Z. Zhang, Y. Liu, L. Liu, Treatment of 5 Critically Ill Patients With COVID-19 With Convalescent Plasma. JAMA, (2020).

4. K. Duan, B. Liu, C. Li, H. Zhang, T. Yu, J. Qu, M. Zhou, L. Chen, S. Meng, Y. Hu, C. Peng, M. Yuan, J. Huang, Z. Wang, J. Yu, X. Gao, D. Wang, X. Yu, L. Li, J. Zhang, X. Wu, B. Li, Y. Xu, W. Chen, Y. Peng, Y. Hu, L. Lin, X. Liu, S. Huang, Z. Zhou, L. Zhang, Y. Wang, Z. Zhang, K. Deng, Z. Xia, Q. Gong, W. Zhang, X. Zheng, Y. Liu, H. Yang, D. Zhou, D. Yu, J. Hou, Z. Shi, S. Chen, Z. Chen, X. Zhang, X. Yang, Effectiveness of convalescent plasma therapy in severe COVID-19 patients. Proc Natl Acad Sci U S A 117, 9490–9496 (2020).

5. M. J. Joyner, R. S. Wright, D. Fairweather, J. W. Senefeld, K. A. Bruno, S. A. Klassen, R. E. Carter, A. M. Klompas, C. C. Wiggins, J. R. Shepherd, R. F. Rea, E. R. Whelan, A. J. Clayburn, M. R. Spiegel, P. W. Johnson, E. R. Lesser, S. E. Baker, K. F. Larson, J. G. Ripoll, K. J. Andersen, D. O. Hodge, K. L. Kunze, M. R. Buras, M. N. Vogt, V. Herasevich, J. J. Dennis, R. J. Regimbal, P. R. Bauer, J. E. Blair, C. M. van Buskirk, J. L. Winters, J. R. Stubbs, N. S. Paneth, N. C. Verdun, P. Marks, A. Casadevall, Early safety indicators of COVID-19 convalescent plasma in 5,000 patients. J Clin Invest, (2020).

6. L. Li, W. Zhang, Y. Hu, X. Tong, S. Zheng, J. Yang, Y. Kong, L. Ren, Q. Wei, H. Mei, C. Hu, C. Tao, R. Yang, J. Wang, Y. Yu, Y. Guo, X. Wu, Z. Xu, L. Zeng, N. Xiong, L. Chen, J. Wang, N. Man, Y. Liu, H. Xu, E. Deng, X. Zhang, C. Li, C. Wang, S. Su, L. Zhang, J. Wang, Y. Wu, Z. Liu, Effect of Convalescent Plasma Therapy on Time to Clinical Improvement in Patients With Severe and Life-threatening COVID-19: A Randomized Clinical Trial. JAMA, (2020).

7. S. T. H. Liu, H.-M. Lin, I. Baine, A. Wajnberg, J. P. Gumprecht, F. Rahman, D. Rodriguez, P. Tandon, A. Bassily-Marcus, J. Bander, C. Sanky, A. Dupper, A. Zheng, D. R. Altman, B. K. Chen, F. Krammer, D. R. Mendu, A. Firpo-Betancourt, M. A. Levin, E. Bagiella, A. Casadevall, C. Cordon-Cardo, J. S. Jhang, S. A. Arinsburg, D. L. Reich, J. A. Aberg, N. M. Bouvier, Convalescent plasma treatment of severe COVID-19: A matched control study. medRxiv, 2020.2005.2020.20102236 (2020).

8. S. J. Zost, P. Gilchuk, J. B. Case, E. Binshtein, R. E. Chen, J. X. Reidy, A. Trivette, R. S. Nargi, R. E. Sutton, N. Suryadevara, L. E. Williamson, E. C. Chen, T. Jones, S. Day, L. Myers, A. O. Hassan, N. M. Kafai, E. S. Winkler, J. M. Fox, J. J. Steinhardt, K. Ren, Y. M. Loo, N. L. Kallewaard, D. R. Martinez, A. Schafer, L. E. Gralinski, R. S. Baric, L. B. Thackray, M. S. Diamond, R. H. Carnahan, J. E. Crowe, Potently neutralizing human antibodies that block SARS-CoV-2 receptor binding and protect animals. bioRxiv, (2020).

9. C. Walls, Y. J. Park, M. A. Tortorici, A. Wall, A. T. McGuire, D. Veesler, Structure, Function, and Antigenicity of the SARS-CoV-2 Spike Glycoprotein. Cell 181, 281–292 e286 (2020).

10. Q. Wang, Y. Zhang, L. Wu, S. Niu, C. Song, Z. Zhang, G. Lu, C. Qiao, Y. Hu, K. Y. Yuen, Q. Wang, H. Zhou, J. Yan, J. Qi, Structural and Functional Basis of SARS-CoV-2 Entry by Using Human ACE2. Cell 181, 894–904 e899 (2020).

11. J. Shang, Y. Wan, C. Luo, G. Ye, Q. Geng, A. Auerbach, F. Li, Cell entry mechanisms of SARS-CoV-2. Proc Natl Acad Sci U S A 117, 11727–11734 (2020).

12. L. A. VanBlargan, L. Goo, T. C. Pierson, Deconstructing the Antiviral Neutralizing-Antibody Response: Implications for Vaccine Development and Immunity. Microbiol Mol Biol Rev 80, 989–1010 (2016).

13. P. J. Klasse, Neutralization of Virus Infectivity by Antibodies: Old Problems in New Perspectives. Adv Biol 2014, (2014).

14. H. A. Vanderven, S. J. Kent, The protective potential of Fc-mediated antibody functions against influenza virus and other viral pathogens. Immunol Cell Biol 98, 253–263 (2020).

15. E. M. Bloch, S. Shoham, A. Casadevall, B. S. Sachais, B. Shaz, J. L. Winters, C. van Buskirk, B. J. Grossman, M. Joyner, J. P. Henderson, A. Pekosz, B. Lau, A. Wesolowski, L. Katz, H. Shan, P. G. Auwaerter, D. Thomas, D. J. Sullivan, N. Paneth, E. Gehrie, S. Spitalnik, E. Hod, L. Pollack, W. T. Nicholson, L. A. Pirofski, J. A. Bailey, A. A. Tobian, Deployment of convalescent plasma for the prevention and treatment of COVID-19. J Clin Invest, (2020).

16. K. Duan, B. Liu, C. Li, H. Zhang, T. Yu, J. Qu, M. Zhou, L. Chen, S. Meng, Y. Hu, C. Peng, M. Yuan, J. Huang, Z. Wang, J. Yu, X. Gao, D. Wang, X. Yu, L. Li, J. Zhang, X. Wu, B. Li, Y. Yu, W. Chen, Y. Peng, Y. Hu, L. Lin, X. Liu, S. Huang, Z. Zhou, L. Zhang, Y. Wang, Z. Zhang, K. Deng, Z. Xia, Q. Gong, W. Zhang, X. Zheng, Y. Liu, H. Yang, D. Zhou, D. Yu, J. Hou, Z. Shi, S. Chen, Z. Chen, X.-x. Zhang, X. Yang, The feasibility of convalescent plasma therapy in severe COVID-19 patients: a pilot study. medRxiv, 2020.2003.2016.20036145 (2020).

17. X. Wang, X. Guo, Q. Xin, Y. Pan, J. Li, Y. Chu, Y. Feng, Q. Wang, Neutralizing Antibodies Responses to SARS-CoV-2 in COVID-19 Inpatients and Convalescent Patients. medRxiv, 2020.2004.2015.20065623 (2020).

18. B. Zhang, X. Zhou, C. Zhu, F. Feng, Y. Qiu, J. Feng, Q. Jia, Q. Song, B. Zhu, J. Wang, Immune phenotyping based on neutrophil-to-lymphocyte ratio and IgG predicts disease severity and outcome for patients with COVID-19. medRxiv, 2020.2003.2012.20035048(2020).

19. E. P. Scully, J. Haverfield, R. L. Ursin, C. Tannenbaum, S. L. Klein, Considering how biological sex impacts immune responses and COVID-19 outcomes. Nat Rev Immunol, (2020).

20. K. L. Flanagan, A. L. Fink, M. Plebanski, S. L. Klein, Sex and Gender Differences in the Outcomes of Vaccination over the Life Course. Annu Rev Cell Dev Biol 33, 577–599 (2017).

21. X. Yu, R. Prados-Rosales, E. R. Jenny-Avital, K. Sosa, A. Casadevall, J. M. Achkar, Comparative evaluation of profiles of antibodies to mycobacterial capsular polysaccharides in tuberculosis patients and controls stratified by HIV status. Clin Vaccine Immunol 19, 198–208 (2012).

22. FDA, FDA, Ed. (2020), vol. 2020.

23. D. Stadlbauer, F. Amanat, V. Chromikova, K. Jiang, S. Strohmeier, G. A. Arunkumar, J. Tan, D. Bhavsar, C. Capuano, E. Kirkpatrick, P. Meade, R. N. Brito, C. Teo, M. McMahon, V. Simon, F. Krammer, SARS-CoV-2 Seroconversion in Humans: A Detailed Protocol for a Serological Assay, Antigen Production, and Test Setup. Curr Protoc Microbiol 57, e100 (2020).

24. S. Matsuyama, N. Nao, K. Shirato, M. Kawase, S. Saito, I. Takayama, N. Nagata, T. Sekizuka, H. Katoh, F. Kato, M. Sakata, M. Tahara, S. Kutsuna, N. Ohmagari, M. Kuroda, T. Suzuki, T. Kageyama, M. Takeda, Enhanced isolation of SARS-CoV-2 by TMPRSS2-expressing cells. Proc Natl Acad Sci U S A 117, 7001–7003 (2020).

25. S. R. Schaecher, E. Touchette, J. Schriewer, R. M. Buller, A. Pekosz, Severe acute respiratory syndrome coronavirus gene 7 products contribute to virus-induced apoptosis. J Virol 81, 11054–11068 (2007).

26. S. R. Schaecher, J. M. Mackenzie, A. Pekosz, The ORF7b protein of severe acute respiratory syndrome coronavirus (SARS-CoV) is expressed in virus-infected cells and incorporated into SARS-CoV particles. J Virol 81, 718–731 (2007).

27. L. J. Reed, H. Meunch, A simple method of estimating 50 percent endpoints. American Journal of Hygiene 27, 493 (1938).

28. Q. Wang, Q. Du, B. Guo, D. Mu, X. Lu, Q. Ma, Y. Guo, L. Fang, B. Zhang, G. Zhang, X. Guo, A Method To Prevent SARS-CoV-2 IgM False Positives in Gold Immunochromatography and Enzyme-Linked Immunosorbent Assays. J Clin Microbiol 58, (2020).

29. S. R. Schaecher, J. Stabenow, C. Oberle, J. Schriewer, R. M. Buller, J. E. Sagartz, A. Pekosz, An immunosuppressed Syrian golden hamster model for SARS-CoV infection. Virology 380, 312–321 (2008).

30. R. Williams, Using the margins command to estimate and interpret adjusted predictions and marginal effects. The Stata Journal 12, 308–331 (2012).

